# The quality of early relational health modifies the effect of early life stress on child emerging psychopathology

**DOI:** 10.64898/2026.01.23.26344636

**Authors:** Jennifer Warmingham, Andréane Lavallée, Paul Curtin, Jill Owen, Katrina Fuller, Hajer Nakua, Hope Hendry, Marissa Lanoff, Angela Gigliotti, Jenna Russo, Vitoria Chaves, Elena Arduin, Nicole Shearman, Imaal Ahmed, Ashley N. Battarbee, Morgan Firestein, Maha Hussain, Margaret Kyle, Rachel Marsh, Alan T. Tita, Michael Varner, Ruiyang Xu, Melissa S. Stockwell, Catherine Monk, Dani Dumitriu

**Affiliations:** Center for Early Relational Health, Columbia University Irving Medical Center, New York, NY; Department of Pediatrics, Vagelos College of Physicians and Surgeons, Columbia University Irving Medical Center, New York, NY; University of Rochester, Mt. Hope Family Center, Rochester, NY; Department of Psychiatry, Columbia University Irving Medical Center, New York, NY; Department of Obstetrics and Gynecology, University of Alabama at Birmingham, Birmingham, AL; Department of Pediatrics and Child Health Institute of New Jersey, Rutgers Robert Wood Johnson Medical School, New Brunswick, NJ; Department of Obstetrics and Gynecology, University of Utah Health Sciences Center, Salt Lake City, UT; New York State Psychiatric Institute, New York, NY; Department of Population and Family Health, Mailman School of Public Health, Columbia University Irving Medical Center, New York, NY; Department of Obstetrics and Gynecology, Columbia University Irving Medical Center, New York, NY

**Keywords:** early life stress, early relational health, child psychopathology, emotional connection

## Abstract

**Background:** Early relational health (ERH) is thought to buffer the association between early life stress (ELS) and child psychopathology, but limited work has directly tested this hypothesis.

**Objective:** We evaluate mother-infant emotional connection, a facet of ERH, as a buffer of combined and individual impacts of specific ELS exposures (maternal mental health and interpartner conflict) on child psychopathology.

**Methods:** Participants included mother-infant dyads (n=100) followed longitudinally in the COMBO cohort, a convenience sample recruited during the COVID-19 pandemic. ERH was assessed via a remote mother-infant face-to-face interaction at ~4mo postpartum coded for emotional connection. An ELS Index was estimated using measures of maternal self-reported postpartum anxiety, depression, stress, and inter-partner conflict. Mothers rated emerging signs of child psychopathology symptoms at 2-3yrs on the Child Behavior Checklist for Ages 1½−5 (CBCL/1½−5). Main and interactive effects of ELS and ERH on emerging signs of child psychopathology were tested in generalized linear models.

**Results:** Greater ELS Index scores were associated with a higher rate of emerging psychopathology symptoms (aRR=1.32, p<.001), but this association was moderated by a significant interaction between the ELS Index and emotional connection (aRR=0.99, p=.03), such that at higher levels of emotional connection, the association of ELS with child psychopathology symptoms was weaker (aRR=1.16, *p*<.001).

**Conclusion:** Parent-infant emotional connection may buffer the impact of ELS exposure in infancy on child emerging symptoms of psychopathology in toddlerhood, supporting efforts to invest in pediatric interventions that target ERH.

## Introduction

Early life stress (ELS) exposures, including parental mental health challenges, violence in the home, death or separation from a caregiver, and various forms of abuse^1^ increase the risk for development of psychopathology in children^2–5^. However, not all children exposed to ELS develop clinically-relevant psychopathology symptoms^6^, likely due to complex interactions between risk and protective factors^7,8^. Early relational health (ERH), or the quality of parent/caregiver-child relationships early in life, is implicated in child socioemotional development and posited to buffer effects of stress^7,9^. Positive ERH can exist in the presence of ELS exposures. Therefore, among children with exposure to adversity, ERH may be one modifiable factor that can mitigate negative effects of stress^7^. Considering the interactive effects of ELS exposures and ERH early in development (i.e., birth to age 3) is critical because infants rely heavily on caregivers to recover from stress^8^. Identifying early intervention/prevention targets to mitigate the severity of psychopathology symptoms associated with ELS is essential to promote health in pediatric populations.

Most work on ERH in the context of ELS focuses on the impact of cumulative^10^ or specific effects^1,11,12^ of stressors or stress patterns^13^ on components of ERH (e.g., disorganized attachment^14^) or focuses on maternal behavior/sensitivity as a buffer of early exposure to family conflict^15–18^. Far less work has investigated dyadic processes that capture the quality of interaction between an infant and caregiver. Dyadic interaction quality is considered an emergent property of the interaction and is therefore more than the “sum of its parts” (e.g., maternal and child behaviors). One dimension of dyadic interaction includes synchrony, a distinct component of ERH relevant for child socioemotional functioning^19,20^ that captures the emotional and biobehavioral dynamics of the parent-infant interaction, rather than focusing strictly on contingent or sensitivity of parental behavior^21,22^. Investigating the buffering effects of this component of ERH is novel and will extend the literature on how dyadic components of ERH function in tandem with ELS^7,23^.

To explore the buffering effect of dyadic emotional connection, we use a machine learning approach to empirically model the co-occurrence and severity of exposure types but also retain information about the individual importance of each exposure type. This approach is poised to extend the literature on ELS^24,25^ and help clarify the effect that ERH may play as a protective factor in the context of complex stress exposures patterns. We further extend the literature by focusing on the dyadic construct of emotional connection,^26^ a so far untested ERH measure as a buffer of ELS on the emergence of psychopathology.

## Methods

### Study Design and Participants

This study used a convenience sample of mother-infant dyads followed longitudinally from infancy to age three who were recruited through the COMBO (previously known as the COVID-19 Mother Infant Outcomes) Initiative, a study of mother and child health in the context of the COVID-19 pandemic. Mother-infant dyads were enrolled either into the original COMBO cohort at Columbia University (New York-based recruitment), or the 3-site (Utah, Alabama, New York) Epidemiology of SARS-CoV-2 in Pregnancy and Infant (ESPI) COMBO sub-study. Mothers with and without maternal prenatal SARS-CoV-2 infection were recruited between May 2020 and April 2025. See **Appendix 1** for detailed information regarding COMBO and ESPI COMBO enrollment.

Participants voluntarily enrolled, consented, completed surveys and video visit assessments, authorized the review of their medical records, and received fiscal compensation for participation. All study procedures were reviewed and approved by the Columbia University Irving Medical Center (CUIMC) Institutional Review Board (IRB), which was the central IRB for data collection sites (CUIMC, University of Utah and University of Alabama at Birmingham IRBs; See 45 C.F.R. part 46.114; 21 C.F.R. part 56.114). Study procedures were completed in English or Spanish. The current study includes a subset of participants who completed all key maternal mental health and inter-partner conflict surveys in the early postpartum period, a video visit with their infants at ~4mo postpartum, and survey ratings of child behavior problems when their children were 2 or 3 years old.

## Measures

### Early Life Stress Exposures

#### Inter-Partner Conflict

The Conflict Tactics Scale^27,28^ (CTS2S) is a 20-item self-report survey completed at ~2 months postpartum that assesses frequency of conflict tactics (e.g., negotiation, types of aggression) used by participants and their romantic partners. Mothers who were currently in a relationship completed this survey. Mothers rated the frequency of each conflict tactic used by themselves and their partner in the last year on this response scale: “Once in the past year”, “Twice in the past year”, “3-5 times in the past year”, “6-10 times in the past year”, “11-20 times in the past year”, “More than 20 times in the past year”, “Not in the past year, but it did happen before”, and “This never happened”. We computed two scores for the frequency of conflict in the past year: one for positive inter-partner conflict (negotiation, compromise, range 0-40), and one for verbal/physical/sexual conflict, which we termed intimate partner violence (IPV, range 0-80).

#### Maternal Postpartum Mental Health

Mothers completed surveys assessing mental health at ~4 months postpartum. **Depressive Symptoms:** The Patient Health Questionnaire (PHQ-9^29,30^) is a self-report measure that assesses depression symptom severity of 9 items (e.g., “Feeling down, depressed, or hopeless”) in the previous two weeks. Response options were 0 (not at all), 1 (several days), 2 (more than half the days), or 3 (nearly every day). Items were summed (range 0-27), with higher total scores indicating greater severity of depression symptoms. **Stress**: The Perceived Stress Scale (PSS^31,32^) is a 14-item instrument that measures the degree to which participants experience events in their lives as stressful. Item scores were summed (range 0-56) with higher total scores indicating greater stress levels. **Anxiety:** The State Anxiety Scale (STAI-S^33,34^) is a 20-item subscale from the State-Trait Anxiety Inventory evaluating the presence and intensity of current anxiety symptoms. Mothers rated how they feel “right now” in response to questions including “I feel tense,” or “I feel nervous.” Scores were summed (range 20-80), with higher total scores indicating greater anxiety.

#### Early Relational Health

The Welch Emotional Connection Screen (WECS^26^) was used to code emotional connection during a 3-minute face-to-face Zoom-recorded interaction between the mother and infant at ~4 months postpartum. Mother-infant videos were coded by reliable research assistants trained in the WECS coding system. Coders assigned scores using a 9-point Likert scale (1-3 in 0.25 increments) on four subscales measuring dyadic interaction quality: Attraction, Vocal Communication, Facial Expressiveness, and Sensitivity/Reciprocity. Subscales were summed (range 4-12), with higher total scores indicating greater emotional connection. Coders double coded 20% of videos and reached an Intraclass correlation (ICC) of 0.98.

#### Early Signs of Child Psychopathology

The Child Behavior Checklist for Ages 1½−5 (CBCL/1½−5^35^) was completed by mothers at 2 and/or 3 years. If the 3-year survey was available we used this timepoint, otherwise the 2-year survey was used. This was done so as not to exclude children born in 2022, who would not have aged into the 3-year assessment at the time that data was pulled for this analysis (Fall of 2024). This widely used instrument includes 99 items relating to behavioral and emotional symptoms with response options including 0-”Not true (as far as you know)”, 1-”Somewhat or sometimes true”, to 2-”Very true or often true.” The Total Problems subscale combines Internalizing and Externalizing subscale scores (range 0-198). Greater scores indicate greater total emotional/behavioral problems.

#### Covariates & Demographic Data

Sociodemographic variables (e.g., maternal age, maternal race, maternal ethnicity, gestational age/prematurity status, language, child year of birth) were assessed using online surveys and/or via medical record extraction.

#### SARS-CoV-2 infection status

Because we used a sample of convenience from a cohort recruited during the COVID-19 pandemic, maternal SARS-CoV-2 infection status during pregnancy was available on all mothers. Detailed assessment methods have been previously published^36,37^ and are included in **Appendix 1**.

### Statistical Analysis

#### Preliminary Analyses

Because the parent study recruited mothers with and without SARS-CoV-2 infection in pregnancy and this recruitment strategy may have impacted our variables of interest, we evaluated differences between mothers with and without SARS-CoV-2 infection in pregnancy on key study variables using t-tests or chi-squared tests prior to evaluating main study aims/analyses.

### Main Analyses

#### Mixtures Modeling: Weighted Quantile Sum (WQS) Regression

To generate a global measure of a child’s ELS exposures, we created an empirically-estimated ELS Index using Weighed Quantile Sum (WQS) regression. WQS is an ensemble-based supervised machine learning technique described in full in prior publications^38–40^ and previously applied to evaluate the impact of fetal and early child exposures on child development^41–43^. The ELS Index is comprised of weighted sum scores of each ELS measure (maternal-reported depression, anxiety, stress, IPV and positive inter-partner conflict) and their relative contribution to CBCL/1½−5 total scores. To estimate the ELS Index itself as well as the individual variable weights within the index, a bootstrap ensemble method was employed (n=100 bootstraps). Higher ELS Index scores indicate greater contribution of individual ELS measures to child psychopathology. The weights for each ELS measure are interpreted as the extent to which that component (e.g., maternal depression) relates to the outcome (CBCL/1½−5 total scores), controlling for other mixture components as well as CBCL/1½−5 model covariates that are *not* in the ELS Index.

Within the WQS regression, we tested main and interactive effects of the ELS Index and emotional connection on CBCL/1½−5 total scores, controlling for child sex and age at CBCL/1½−5 assessment. Given that CBCL/1½−5 scores are distributed on a non-negative integer scale, Poisson models were used in effect estimation. If the interaction term was significant (p<0.05), we interpreted the association between the ELS Index and child psychopathology to be significantly

#### Univariate Multivariate Regression Models

We also conducted analyses evaluating whether emotional connection buffers each ELS component individually by testing main and interactive effects of each ELS measure and emotional connection on CBCL/1½−5 total scores in five separate generalized linear models (with Poisson distribution), controlling for child sex and age at CBCL/1½−5 assessment. Simple slopes analyses were conducted as described above if the interaction term was significant (*p*<0.05).

For all regression models, adjusted rate ratios [aRR] were computed. All analyses were conducted in R version 4.3.1^44^. We used the *gWQS (v3*.*0*.*5)* package to estimate the WQS regression model and *interactions (v1*.*2*.*0)* and *ggplot2 (v3*.*5*.*1)* packages to test simple slopes and generate interactions plots.

## Results

### Participants & Descriptive Statistics

Participants included n=100 mother-child dyads who had complete data regarding: (1) inter-partner conflict (CTS2S), (2) maternal postpartum mental health (PHQ-9, STAI-S, PSS), (3) emotional connection, and (4) CBCL/1½−5. Children (42.0% female) were born between 6/2020-9/2022. This subset of mothers self-identified as 31.0% Hispanic, 54.0% White, 10.0% Black, 7.0% Asian, 18.0% Other/More than one category, and 11.0% unknown race/ethnicity and included participants recruited from either the NY (n=65, 65.0%) or the Utah (n=35, 35.0%), as no Alabama-based dyad had complete data. Of those included, n=78 (78.0%) completed study procedures in English and n=22 (22.0%) in Spanish. Additional demographic data are presented in **Table 1**.

**Table 1.** Demographics.

|  |  | **Total** | **NY** | **UT** |
| --- | --- | --- | --- | --- |
|  |  | n=100 | n=65 | n=35 |
|  |  | N (%) | N (%) | N (%) |
| Child year of birth | 2020 | 29 (29.0) | 29 (44.6) | 0 (0) |
|  | 2021 | 67 (67.0) | 32 (49.2) | 35 (100.0) |
|  | 2022 | 4 (4.0) | 4 (6.2) | 0 (0) |
| Maternal language | English | 78 (78.0) | 43 (66.2) | 35 (100.0) |
|  | Spanish | 22 (22.0) | 22 (33.8) | 0 (0) |
| Insurance | Commercial | 65 (65.0) | 33 (50.8) | 32 (91.4) |
|  | Medicaid | 35 (35.0) | 32 (49.2) | 3 (8.6) |
| Pregnancy SARS-CoV-2 Status | Detected (Positive) | 25 (25.0) | 14 (21.5) | 11 (31.4) |
|  | Not Detected (Negative) | 75 (75.0) | 51 (78.5) | 24 (68.6) |
| Preterm Birth | No | 99 (99.0) | 65 (100.0) | 34 (97.1) |
|  | Yes | 1 (1.0) | 0 (0) | 1 (2.9) |
| Child sex | Female | 42 (42.0) | 22 (33.8) | 20 (57.1) |
|  | Male | 58 (58.0) | 43 (66.2) | 15 (42.9) |
| Maternal Age at Delivery | Mean (SD) | 32.1 (5.7) | 33.2 (5.4) | 30.2 (5.8) |
| Maternal Race | Asian | 7 (7.0) | 7 (10.8) | 0 (0) |
|  | Black/African-American | 10 (10.0) | 10 (15.4) | 0 (0) |
|  | White | 54 (54.0) | 20 (30.8) | 34 (97.1) |
|  | More than One Category | 6 (6.0) | 5 (7.7) | 1 (2.9) |
|  | Other | 12 (12.0) | 12 (18.5) | 0 (0) |
|  | I don't know/Prefer Not to Answer | 11 (11.0) | 11 (16.9) | 0 (0) |
| Maternal Ethnicity | Hispanic | 31 (31.0) | 29 (44.6) | 2 (5.7) |
|  | Not Hispanic | 64 (64.0) | 31 (47.7) | 33 (94.3) |
|  | I don't know/prefer not to answer | 5 (5.0) | 5 (7.7) | 0 (0) |
| Infant age 4mo video visit | Mean (SD) months | 4.7 (1.1) | 4.6 (0.8) | 4.8 (1.5) |

Note: NY = New York, UT = Utah Study Sites different across levels of emotional connection and simple slope analyses were conducted to evaluate the association between the ELS Index and child psychopathology symptoms at different levels (+1SD and -1SD) of emotional connection.

Data were drawn from a convenience sample of mother-infant dyads enrolled during the COVID-19 pandemic, so we first explored potential SARS-CoV-2 infection status as a confounder. We found no association between SARS-CoV-2 status in pregnancy and key study variables (maternal mental health, CBCL/1½−5 Total Problems score, inter-partner conflict, emotional connection; *p*s=0.2-0.9; See Appendix 2 for sample means on key variables and means within SARS-CoV-2 status).

### Mother-infant emotional connection modifies the association between a global measure of ELS and future child psychopathology

Maternal depression and anxiety symptoms most strongly contributed to the ELS Index (**Figure 1A**). Greater scores on the ELS Index were associated with higher rates of CBCL Total Problems scores (adjusted rate ratio [aRR]=1.32, 95% CI[1.20–1.45]), a main effect that was modified by a significant interaction between the ELS Index and emotional connection (aRR = 0.99, 95% CI [0.97-.99]) (**Figure 1B**). This suggests that the association between the ELS Index and CBCL Total Problems scores depends on, and should be interpreted in the context of, the level of emotional connection.

**Figure 1.**
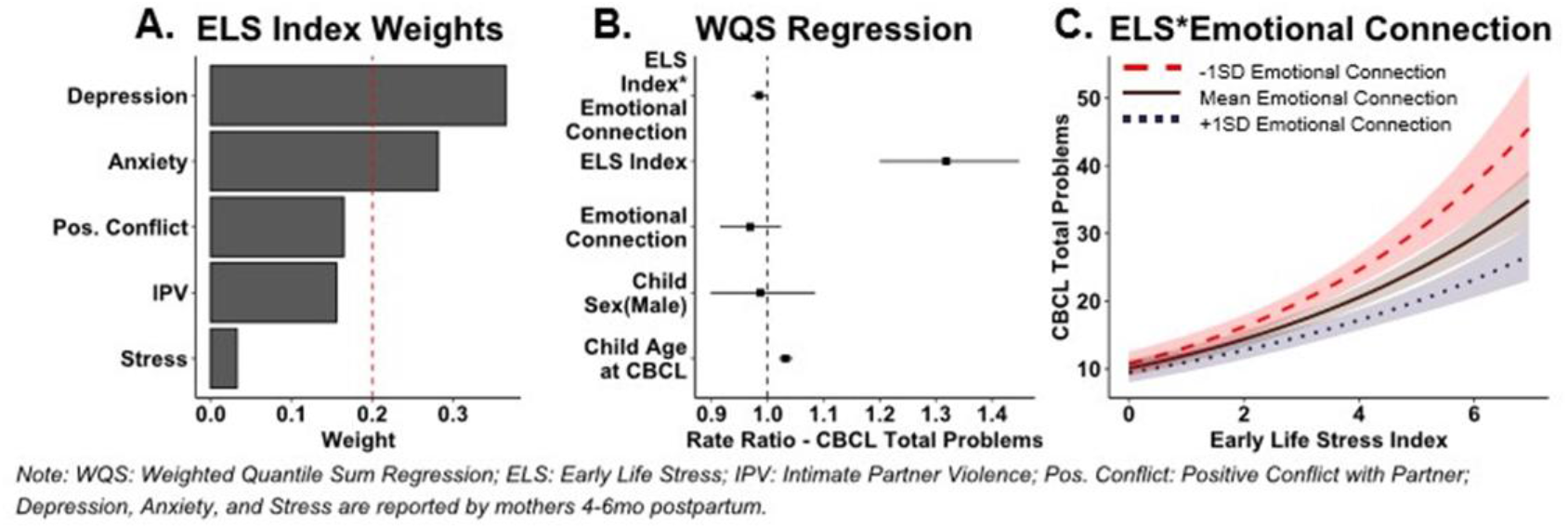
Impact of ELS and Emotional Connection on Child Psychopathology.

To visualize this interaction, we plotted the effect of the ELS Index on rates of CBCL Total Problems scores at different levels of emotional connection: at the mean, high levels of emotional connection (+1SD) and low levels of emotional connection (−1SD). At all levels of emotional connection, higher scores of the ELS Index was associated with greater rates of CBCL Total Problems scores, but this effect was weaker at higher levels of emotional connection (**Figure 1C**). When emotional connection scores were low (−1SD[4.49]), a one point increase on the ELS Index was associated with a 23% increase in the rate of CBCL Total Problems scores (aRR=1.23, *p*<.001). Comparatively, when emotional connection scores were high (+1SD, [8.40]), a one point increase on the ELS Index was associated with only a 16% increase in the rate of CBCL Total Problems scores (aRR=1.16, *p*<.001; **Figure 1C**).

### Mother-infant emotional connection selectively buffers specific types of ELS

Maternal depression, anxiety, stress, and IPV in infancy were each associated with increased rates of CBCL Total Problems scores (left panels of **Figures 2A, 2B, 2C**, and **2D**). These effects did not depend on (interact with) emotional connection. A one point increase in maternal stress was associated with a 2% increase in the rate of CBCL Total Problems scores (**Figure 2C**; aRR=1.02, 95% CI[1.00–1.04]). For maternal anxiety, a one point increase was assoicated with a 4% increase the rate of CBCL Total Problems scores (**Figure 2B** left panel; aRR=1.04, 95% CI [1.02–1.05]). Positive inter-partner conflict was not associated with CBCL Total Problems scores (**Figure 2E**, left panel).

**Figure 2.**
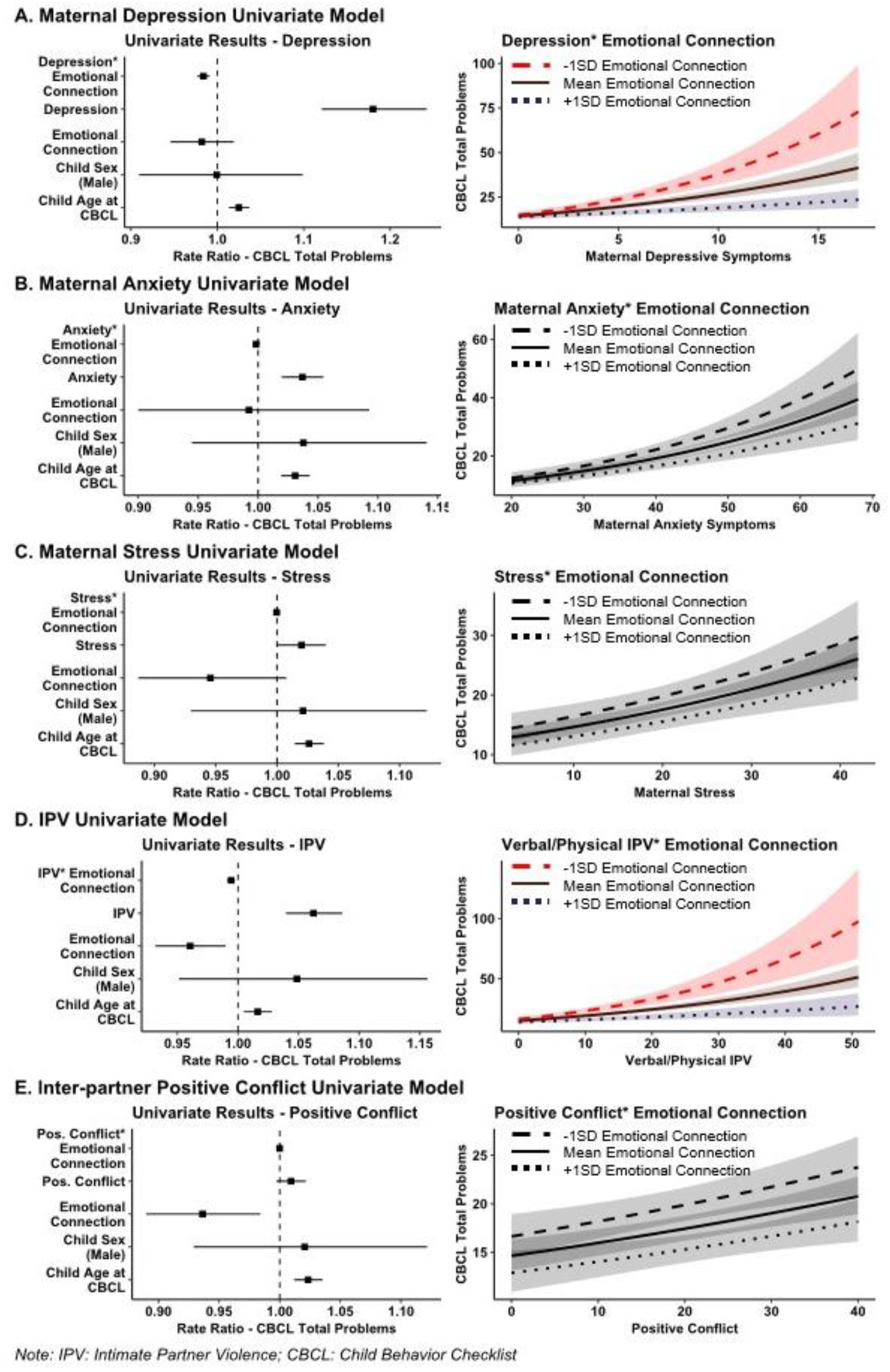
Univariate Models. Note: IPV: Intimate Partner Violence; CBCL Child Behavior Checklist

Mother-infant emotional connection moderated the effect of maternal depression (**Figure 2A**; depression main effect: aRR=1.18, 95% CI[1.12–1.24]; depression^*^emotional connection interaction: aRR=1.18, 95% CI [0.98–0.99]) and IPV (**Figure 2D**; IPV main effect: aRR=1.06, 95% CI[1.04–1.09]; emotional connection^*^IPV interaction: aRR=0.99, 95% CI[0.99–1.00]) on rate of CBCL Total Problems scores. At low levels of emotional connection, a one point increase in maternal depression scores was associated with a 9% increase in the rate of CBCL Total Problems scores (aRR=1.09, *p*<.001), but at high levels of emotional connection a one point increase in maternal depression scores associated with only 3% increase in the rate of CBCL Total Problems scores (aRR=1.03, *p*<.001) (**Figure 2A**, right panel). Similarly, at low levels of emotional connection, a one point increase in IPV frequency was associated with a 3% increase in the rate of CBCL Total Problems scores (aRR=1.03, *p*<.001). At high levels of emotional connection, an one point increase in IPV resulted in only 1% increase in the rate of CBCL Total Problems scores (aRR=1.01, *p*<.001) (**Figure 2D**, right panel).

## Discussion

The goal of this study was to evaluate observed mother-infant emotional connection, a component of ERH measured in infancy, as a buffer of the impact of ELS exposures on emerging symptoms of psychopathology in toddlerhood, as measured by the CBCL/1.5-5. We evaluated emotional connection as a buffer of both combined and individual ELS exposures (maternal postpartum psychopathology and inter-partner conflict) on child psychopathology. We found that the strength of mother-infant emotional connection buffered the impact of ELS on emerging symptoms of child psychopathology, both at the level of the global empirically-estimated ELS Index, and for two individual exposures: maternal postpartum depression and perinatal intimate partner violence. At lower levels of emotional connection, ELS was more strongly associated with emerging child psychopathology. At higher levels of emotional connection, the impact of ELS was weaker.

Regardless of emotional connection score, greater ELS exposure predicted more early signs of emerging signs of psychopathology, suggesting that ERH may buffer, but not completely counteract the negative impact of ELS on child socioemotional health. This is consistent with work suggesting that postpartum maternal mental health and IPV exposure matters for child health in both low- and high-risk samples^2,3,45–48^. Efforts to prevent diverse and co-occurring forms of ELS^49^ are critical to prevent the development of psychopathology.

Here, we investigated emotional connection^26,50^, a relatively understudied ERH component as compared to attachment or maternal sensitivity^16,18^, as a protective factor in the context of ELS. This study adds to a growing body of work suggesting dyadic synchrony processes – and not just maternal behavior^15,16^ – are buffers of stress^21,22^. Additionally, the combined stressor characterization using an empirically-estimated ELS index may address limitations of approaches to adversity research that consider single exposure types, which do not capture the nuanced co-occurring stress exposures in the lives of children and families^24,25,51^.

Methodological strengths of this study include longitudinal design and rigorous method of characterizing observational emotional connection from a quick interaction between mothers and their infants that can be conducted remotely^26,50^. This type of low participant burden assessment of ERH is essential to advancing scientific knowledge regarding the direct or indirect (buffering) effects of ERH on child outcomes to rapidly inform much-needed evidence-based prevention/intervention targets in pediatric populations^7^. Generalizability of this study may be limited because the data were collected during the COVID-19 pandemic. Another limitation is the relatively low endorsement of early signs of child psychopathology in our sample. This is a relatively small sample, and we have conducted exploratory analyses that were uncorrected for multiple tests, and so we view our results as preliminary and in need of replication in independent samples. Future studies are needed to test similar buffering models in larger samples with different co-occurring types of ELS known to increase the risk for emerging psychopathology (e.g., abuse/neglect, loss, family instability, medical trauma^1,52,53^) to test the boundaries of ERH components as buffers.

## Conclusion

Investigating the buffering effects of ERH on ELS is essential to inform primary care interventions^7^. This preliminary evidence shows that emotional connection, an observational measure of ERH that can be assessed with high fidelity in a short video visit, buffered the effects of certain ELS exposures on emerging signs of child psychopathology. Future replication of these results in an independent sample is essential to establish emotional connection as a tenable buffer of ELS and to guide evidence-based intervention and prevention efforts geared toward the promotion of ERH.

## Data Availability

The datasets analyzed during the current study are available from the corresponding author on reasonable request and pending IRB approval

