## Supplement for "The quality of early relational health modifies the effect of early life stress on child emerging psychopathology"

### Appendix 1. Study Cohort Descriptions

#### COVID-19 Mother-Baby Outcomes (COMBO) Initiative Cohort

The COMBO Initiative recruited women who receive prenatal care and delivered at the Columbia University Irving Medical Center (CUIMC)-affiliated NewYork-Presbyterian (NYP) Morgan Stanley Children's Hospital (MSCH) or NYP Allen Pavilion Hospital. Mothers could also self-refer. Recruitment and enrollment for the COMBO Initiative began in May 2020 and ended in April 2025.

*SARS-CoV-2 Exposed Group:* Pregnant women with positive SARS-CoV-2 testing results during pregnancy (exposed group) where invited to participate. Women with a history of SARS-CoV-2 infection during pregnancy were also enrolled postnatally. Universal nasopharyngeal PCR testing and universal serological testing for SARS-CoV-2 antibodies was implemented by CUIMC for all delivering mothers between March 22, 2020, and July 20, 2020, respectively. For infants born before November 1, 2020, prenatal exposure was determined if the mother had a positive SARS-CoV- 2 PCR and/or serology test documented in the electronic health record during pregnancy or at delivery. After November 1, 2020, positive SARS-CoV-2 status was determined by a positive PCR or antigen test during pregnancy or a positive serology test and documentation of COVID-19 symptoms in the medical record (determined via electronic chart review).

*SARS-CoV-2 Unexposed Group:* Following delivery, each infant born to a mother with a confirmed SARS-CoV-2 infection during pregnancy was matched to one to three infants without documented exposure to a maternal SARS-CoV-2 infection during pregnancy (unexposed group). For infants born before July 20, 2020, the unexposed classification required the absence of PCR positivity in the EHR, no self-reported PCR positivity, absence of COVID symptoms recorded in the EHR and no self-reported COVID symptoms. After July 20, 2020, the unexposed group classification required the absence of PCR positivity and/or a negative serology. Exposed and unexposed groups were matched based on sex, gestational age at birth, mode of delivery, and birthdate within a two-week window.

This study was reviewed and approved by the CUIMC Institutional Review Board (IRB). Mothers signed electronic consent for study procedures and electronic health record review. Enrolled mothers completed study procedures (video visits, surveys) that could be completed in pregnancy until 4 years postpartum that were conducted in English and Spanish. Not all enrolled mothers completed all study procedures. However, enrolled mothers were offered the opportunity to complete study activities prospectively as children aged into survey and/or video visit timepoints.

#### ESPI COMBO Cohort

We also draw data from the CDC-funded Epidemiology of Severe Acute Respiratory Syndrome Coronavirus-2 in Pregnancy and Infancy (ESPI) COMBO sub-study. The ESPI study was a prospective SARS-CoV-2 infection surveillance study that enrolled pregnant women from three academic medical centers in the U.S. (CUIMC, University of Alabama-Birmingham, and

University of Utah)[54]. Women were eligible if they were pregnant and at <28 weeks gestation; aged 18–50 years; willing to self-collect and mail mid-turbinate nasal swab specimens and respond to weekly surveillance contacts; willing to have up to three blood draws during pregnancy/postpartum; willing to have data collected from their infants' medical records at delivery; and able to speak and read either English or Spanish. Women were ineligible if they were enrolled in a COVID-19 or influenza vaccine clinical trial or intended to enroll in a trial during the current pregnancy. Participants self-collected and submitted weekly mid-turbinate nasal swabs for SARS-CoV-2 reverse transcription polymerase chain reaction (RT-PCR) testing, completed weekly questionnaires about illness symptoms, and submitted additional mid-turbinate swabs when experiencing COVID-19–like symptoms. Serum was collected at enrollment, at the end of the second trimester and at end of pregnancy for testing for SARS-CoV-2 antibodies.

Classification of perinatal SARS-CoV-2 status in this study was based on a combination of maternal self-report of COVID prior to ESPI enrollment, PCR testing and symptom report during the ESPI phase, and serological testing during the ESPI phase.

Serological testing in ESPI was conducted from sera collected up to three times during pregnancy (at enrollment, at end of second trimester, and at end of pregnancy). All participants had serological testing performed at the end of pregnancy. Samples were processed at the CDC using Luminex xMAP-SARS-CoV-2 Multi Antigen Assay. The assay tests 3 antigens: S1, RBD, and nucleocapsid (N) proteins. The N protein is unique to natural infection. S1 and RBD antibodies form in response to both natural infection and vaccination. Considering all three antibodies allows for differentiation between positive testing in response to vaccination versus natural infection. The ESPI sample contained individuals positive for S1 and/or RBD (negative for N), who had never been vaccinated, which was attributed to quicker waning of antibodies against N than of antibodies against S1 and RBD.

The following classification considering self-report, PCR results, and antibody positivity was created:

1. **SARS-CoV-2 not detected**
    a. No self-reported COVID <u>and</u>
    b. Absent positive PCR from ESPI enrollment to delivery <u>and</u>
    c. Negative S1, RBD, N on all serological testing in ESPI <u>and</u>
    d. Serology testing was obtained at the end of pregnancy and was negative
2. **SARS-CoV-2 detected: Pre-pregnancy infection with no in utero exposure**
    a. Self-reported COVID infection with a specific date prior to calculated conception date <u>and</u>
    b. Serology consistent with prior infection
        i. Positive N at entry into ESPI <u>or</u>
        ii. Positive RBD/S1 at entry into ESPI and no history of vaccination
3. **SARS-CoV-2 detected: Unknown timing, possible in utero exposure**
    a. No self-report of COVID history, but:

        i. Positive N at entry into ESPI or
        ii. Positive RBD/S1 and no history of vaccination

4. **SARS-CoV-2 detected during pregnancy**
    a. PCR+ from ESPI enrollment to delivery or
    b. Serological conversion from ESPI enrollment to delivery that cannot be explained by vaccination:
        i. Serological conversion with N pos irrespective of vaccine and symptoms or
        ii. Serological conversion with RBD/S1 prior to vaccine irrespective of symptoms

*ESPI COMBO Sub-study*

The ESPI COMBO sub-study recruited ESPI participants who had completed >40% of weekly ESPI surveillance procedures. Mothers enrolled in ESPI COMBO were invited to complete additional online surveys and remote video visits (via Zoom) with their infants until 6 months postpartum. A total of 453 dyads (out of 689 approached) enrolled in the ESPI COMBO study. Although all mothers were invited to complete all ESPI COMBO study procedures, all study procedures were voluntary and therefore some mothers completed more study activities than others. Study procedures (video visits, surveys) were conducted in English and Spanish. This study was reviewed and approved by the CUIMC Institutional Review Board (IRB), which was the central IRB for data collection sites (CUIMC, University of Utah and University of Alabama IRBs) (See 45 C.F.R. part 46.114; 21 C.F.R. part 56.114). The Centers for Disease Control and Prevention relied on the review of the CUIMC IRB. Written informed consent was completed by mothers prior to engagement in both ESPI and ESPI COMBO study procedures, including medical records abstraction. At the end of their participation in ESPI COMBO, mothers could self-refer and enroll into the main COMBO study, and ~350 of 453 dyads entered the main COMBO study.

### Appendix 2. Maternal SARS-CoV-2 Status and Key Study Variables

Table B.1. Mean differences between those with and without SARS-CoV-2 in pregnancy on key study variables.

| | | | Pregnancy SARS-CoV-2 Status | | |
|---|---|---|---|---|---|
| | | Total N=100 | Negative N=75 | Positive N=25 | *p*-value |
| IPV(Physical/Verbal) | Mean (SD) | 4.8 (9.0) | 5.0 (8.5) | 4.3 (10.6) | 0.75 |
| Positive Conflict | Mean (SD) | 20.5 (13.8) | 20.7 (13.8) | 20.1 (14.2) | 0.84 |
| Maternal Depression | Mean (SD) | 3.4 (3.4) | 3.5 (3.3) | 3.4 (3.7) | 0.91 |
| Maternal Anxiety | Mean (SD) | 35.0 (10.3) | 35.3 (10.2) | 34.1 (10.6) | 0.60 |
| Maternal Stress | Mean (SD) | 19.6 (8.9) | 19.7 (8.7) | 19.3 (9.7) | 0.87 |
| CBCL/1½−5 Total Scores | Mean (SD) | 18.0 (16.0) | 19.1 (16.8) | 14.6 (13.2) | 0.22 |
| Emotional Connection | Mean (SD) | 6.4 (2.0) | 6.3 (1.8) | 6.8 (2.4) | 0.29 |

IPV: Physical/Verbal Intimate Partner Violence; CBCL/1½−5: Child Behavior Checklist
